# Reportable range of quantitative assays for SARS-CoV-2 antibodies determination. An overlooked issue?

**DOI:** 10.1101/2021.03.21.21254043

**Authors:** Ruggero Dittadi, Isabella Bertoli, Paolo Carraro

**Affiliations:** Laboratory Medicine Unit, Ospedale dell’Angelo, ULSS 3 Serenissima, Mestre, Italy

**Keywords:** SARS-CoV-2 antibodies, reportable range, dilution, chemiluminescence assay, ELISA

## Abstract

In order to identify the quantization capability of two methods for SARS-CoV-2 IgG determination (Anti-SARS-CoV-2 QuantiVac ELISA IgG from Euroimmun and SARS-CoV-2 IgG anti-RBD from SNIBE), the linearity of the reportable range with respect to the calibration curve was evaluated.

---

The claimed reportable ranges are 6-120 RU/mL for Euroimmun method (positivity threshold 11 RU/mL) and 0.18-100 AU/mL for SNIBE method (positivity threshold 1 AU/mL). 29 serum specimens with different degrees of positivity from both patients recovered from COVID-19 and vaccinated subjects were evaluated (nine for SNIBE method and 20 for Euroimmun method). The results showed that both the methods could satisfactorily quantify anti SARS-CoV-2 antibodies, provided that the highest detectable value is fixed to 50 AU/mL for SNIBE and 100 RU/mL for ELISA. Beyond these concentrations, the samples should be diluted.

There are some evidences showing that anti-SARS-CoV-2 antibody concentrations differ according to the severity of the disease (1,2). Even in the context of assessing the strength and duration of the antibody response, both after illness and after vaccination, the quantization of antibody concentrations may have a significant role. Most of the commercial methods were originally produced with the aim of identifying the positivity for the presence of antibodies at a qualitative level, and it was possible to propose a real quantization only with the introduction of methods with appropriate calibration curves. One way to identify the quantization capability of an immunoassay method is to evaluate the linearity of the reportable range of measurement with respect to the calibration curve (3). This evaluation was carried out for methods Anti-SARS-CoV-2 QuantiVac ELISA IgG (Euroimmun, Lubeck, Germany) and the recently released SARS-CoV-2 IgG anti-RBD (SNIBE, Shenzen, China).

Both the assays were performed according the manufacturer’s instructions. In the ELISA Euroimmun method patients’ samples were prediluted 1:100 with sample buffer, and the concentrations were determined through the interpolation with a six-point calibration curve (from 1 to 120 Relative Units/mL). The SNIBE method were carried out on the analyzer Maglumi 800 (SNIBE; Shenzen, China), and the samples concentrations were determined through the interpolation with a nine-point master curve (from 0.5 to 100 Arbitrary Unit/mL) periodically adjusted by a 2-point calibration.

The claimed reportable ranges are 6-120 RU/mL for Euroimmun method (positivity threshold 11 RU/mL) and 0.18-100 AU/mL for SNIBE method (positivity threshold 1 AU/mL).

29 serum specimens with different degrees of positivity from both patients recovered from COVID-19 and vaccinated subjects were evaluated. Nine of these specimens were used for the dilution tests with the SNIBE method, while 20 specimens were used with the Euroimmun method. The dilutions were carried out using the respective diluents supplied by the companies. The linearity of the dilution was evaluated according to the method suggested by Emerson et al (4) using ± 3 zeta-score as the limit of acceptability. The results are shown in table 1. For the ELISA method, the data resulted below 3 z-scores and the recovery resulted below ± 20% of the initial value in all samples with initial values <100 RU/mL. Among the 6 cases with initial values between 100 and 120 RU/mL, 4 showed a non-linear trend, with subsequent dilutions showing final results constant but higher than the undiluted value (tab.1). For the SNIBE method, an underestimation of values higher than 50 AU/mL was detected, while for concentrations lower than 50 AU/mL the linearity seems confirmed (tab.2). Since the SNIBE method does not provide the predilution of the samples, two samples were also diluted using a negative plasma sample, to maintain the same matrix over the dilution curve. The results were similar to those found using the diluent of the kit (tab.2).

**Table 1.** Dilution test with Euroimmun method.*The z-score was calculated according Emerson et al (4), and considered acceptable up to 3s. The dilution factor “1” denoted the first prediluted patient sample (1/100). The asterisk denoted the non-linear sample dilution*.

| Sample | Dilution factor | Measured value | Final value | Z score | Recovery % |
| --- | --- | --- | --- | --- | --- |
| 1 | 1 | 79,9 | 79,9 | - | - |
|  | 2 | 45,2 | 90,3 | 2,13 | 113,1 |
|  | 4 | 21,4 | 85,6 | 1,19 | 107,1 |
|  | 8 | 11,1 | 88,6 | 1,57 | 110,9 |
| 2 | 1 | 111,6 | 111,6 | - | - |
|  | 2 | 64,2 | 128,5 | 2,48 | 115,1 |
|  | 4 | 31,0 | 124,0 | 1,84 | 111,2 |
|  | 8 | 14,6 | 116,5 | 0,66 | 104,4 |
| 3 | 1 | 78,9 | 78,9 | - | - |
|  | 2 | 36,8 | 73,5 | -1,25 | 93,2 |
|  | 4 | 17,3 | 69,3 | -2,27 | 87,8 |
|  | 8 | 9,1 | 72,9 | -1,24 | 92,4 |
| 4 | 1 | 112,8 * | 112,8 | - | - |
|  | 2 | 68,0 | 135,9 | 3,21 | 120,4 |
|  | 4 | 37,0 | 147,8 | 4,61 | 131,0 |
|  | 8 | 17,8 | 142,5 | 3,47 | 126,3 |
| 5 | 1 | 105,7 * | 105,7 | - | - |
|  | 2 | 72,0 | 144,0 | 5,25 | 136,2 |
|  | 4 | 41,6 | 166,3 | 7,08 | 157,3 |
|  | 8 | 19,8 | 158,0 | 5,16 | 149,5 |
| 6 | 1 | 65,4 | 65,4 | - | - |
|  | 2 | 35,0 | 69,9 | 1,15 | 106,9 |
|  | 4 | 16,6 | 66,3 | 0,23 | 101,4 |
|  | 8 | 8,7 | 70,0 | 1,03 | 107,0 |
| 7 | 1 | 68,6 | 68,6 | - | - |
|  | 2 | 38,8 | 77,7 | 2,15 | 113,2 |
|  | 4 | 17,6 | 70,3 | 0,41 | 102,5 |
|  | 8 | 9,2 | 73,6 | 0,98 | 107,2 |
| 8 | 1 | 75,5 | 75,5 | - | - |
|  | 2 | 42,5 | 84,9 | 2,05 | 112,6 |
|  | 4 | 19,1 | 76,6 | 0,25 | 101,5 |
|  | 8 | 9,6 | 77,0 | 0,28 | 102,0 |
| 9 | 1 | 99,6 | 99,6 | - | - |
|  | 2 | 51,1 | 102,1 | 0,44 | 102,5 |
|  | 4 | 24,6 | 98,5 | -0,19 | 98,9 |
|  | 8 | 13,2 | 105,5 | 0,81 | 105,9 |
| 10 | 1 | 86,0 | 86,0 | - | - |
|  | 2 | 46,1 | 92,2 | 1,20 | 107,2 |
|  | 4 | 22,8 | 91,3 | 1,04 | 106,2 |
|  | 8 | 12,3 | 98,3 | 2,02 | 114,3 |
| 11 | 1 | 96,5 | 96,5 | - | - |
|  | 2 | 48,7 | 97,3 | 0,14 | 100,8 |
|  | 4 | 22,6 | 90,6 | -1,11 | 93,8 |
|  | 8 | 12,1 | 96,7 | 0,02 | 100,1 |
| 12 | 1 | 78,8 | 78,8 | - | - |
|  | 2 | 42,7 | 85,3 | 1,38 | 108,2 |
|  | 4 | 20,0 | 79,9 | 0,24 | 101,4 |
|  | 8 | 10,2 | 81,7 | 0,56 | 103,7 |
| 13 | 1 | 99,0 | 99,0 | - | - |
|  | 2 | 44,7 | 89,4 | -1,77 | 90,3 |
|  | 4 | 21,3 | 85,0 | -2,64 | 85,9 |
|  | 8 | 11,2 | 89,2 | -1,63 | 90,1 |
| 14 | 1 | 70,4 | 70,4 | - | - |
|  | 2 | 34,3 | 68,6 | -0,45 | 97,5 |
|  | 4 | 16,4 | 65,8 | -1,18 | 93,5 |
|  | 8 | 8,6 | 68,5 | -0,42 | 97,3 |
| 15 | 1 | 108,3 | 108,3 | - | - |
|  | 2 | 59,3 | 118,7 | 1,59 | 109,6 |
|  | 4 | 29,2 | 116,8 | 1,31 | 107,8 |
|  | 8 | 14,6 | 116,4 | 1,11 | 107,5 |
| 16 | 1 | 89,9 | 89,9 | - | - |
|  | 2 | 43,9 | 87,9 | -0,41 | 97,7 |
|  | 4 | 21,4 | 85,6 | -0,86 | 95,2 |
|  | 8 | 10,4 | 83,4 | -1,18 | 92,7 |
| 17 | 1 | 76,9 | 76,9 | - | - |
|  | 2 | 41,4 | 82,8 | 1,28 | 107,6 |
|  | 4 | 20,5 | 81,9 | 1,08 | 106,4 |
|  | 8 | 9,5 | 76,0 | -0,19 | 98,8 |
| 18 | 1 | 113,2 * | 113,2 | - | - |
|  | 2 | 70,6 | 141,2 | 3,80 | 124,8 |
|  | 4 | 37,3 | 149,3 | 4,73 | 131,9 |
|  | 8 | 18,9 | 151,1 | 4,24 | 133,5 |
| 19 | 1 | 93,0 | 93,0 | - | - |
|  | 2 | 48,5 | 96,9 | 0,71 | 104,2 |
|  | 4 | 24,3 | 97,1 | 0,75 | 104,4 |
|  | 8 | 11,3 | 90,4 | -0,45 | 97,1 |
| 20 | 1 | 117,2 * | 117,2 | - | - |
|  | 2 | 68,2 | 136,4 | 2,62 | 116,4 |
|  | 4 | 35,2 | 140,7 | 3,14 | 120,0 |
|  | 8 | 17,4 | 139,4 | 2,60 | 118,9 |

Both the assays evaluated use a wide range standard curve, that provide an accurate quantification over the measurement range. However, it is not warranted that high concentrations of detectable antibodies could not saturate the analytical system before to reach the highest calibration point.

An evaluation of dilution curves showed that the QuantiVac-ELISA from Euroimmun and the IgG anti-RBD from SNIBE could satisfactorily quantify anti SARS-CoV-2 antibodies, provided that the highest detectable value is fixed to 50 AU/mL for SNIBE and 100 RU/mL for ELISA. Beyond these concentrations, the samples should be diluted for a more correct quantitation.

## Data Availability

The data referred in the manuscript are available.

## Conflicts of Interest

*No potential conflict of interest was reported by the authors*.

*The data was collected during clinical practice, and the Ethics Committee stated that the review was not required*

## Acknowledgments

*The authors would like to thank Euroimmun and Medical Systems S.p.A. for kindly providing the necessary reagents*.

## Funding

*This research received no specific grant from any funding agency in the public, commercial, or not-for-profit sectors*.

---

**Table 2.**
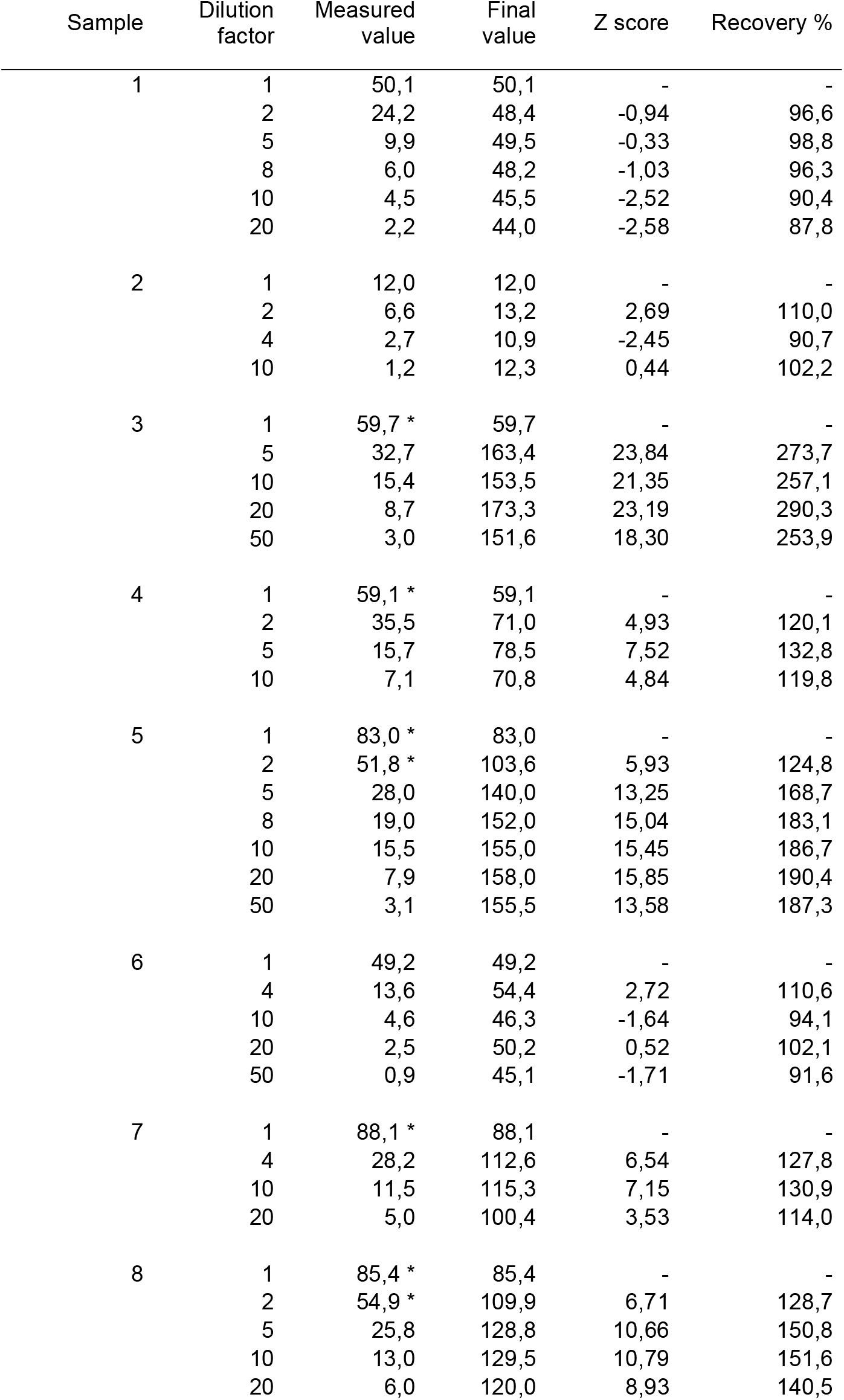
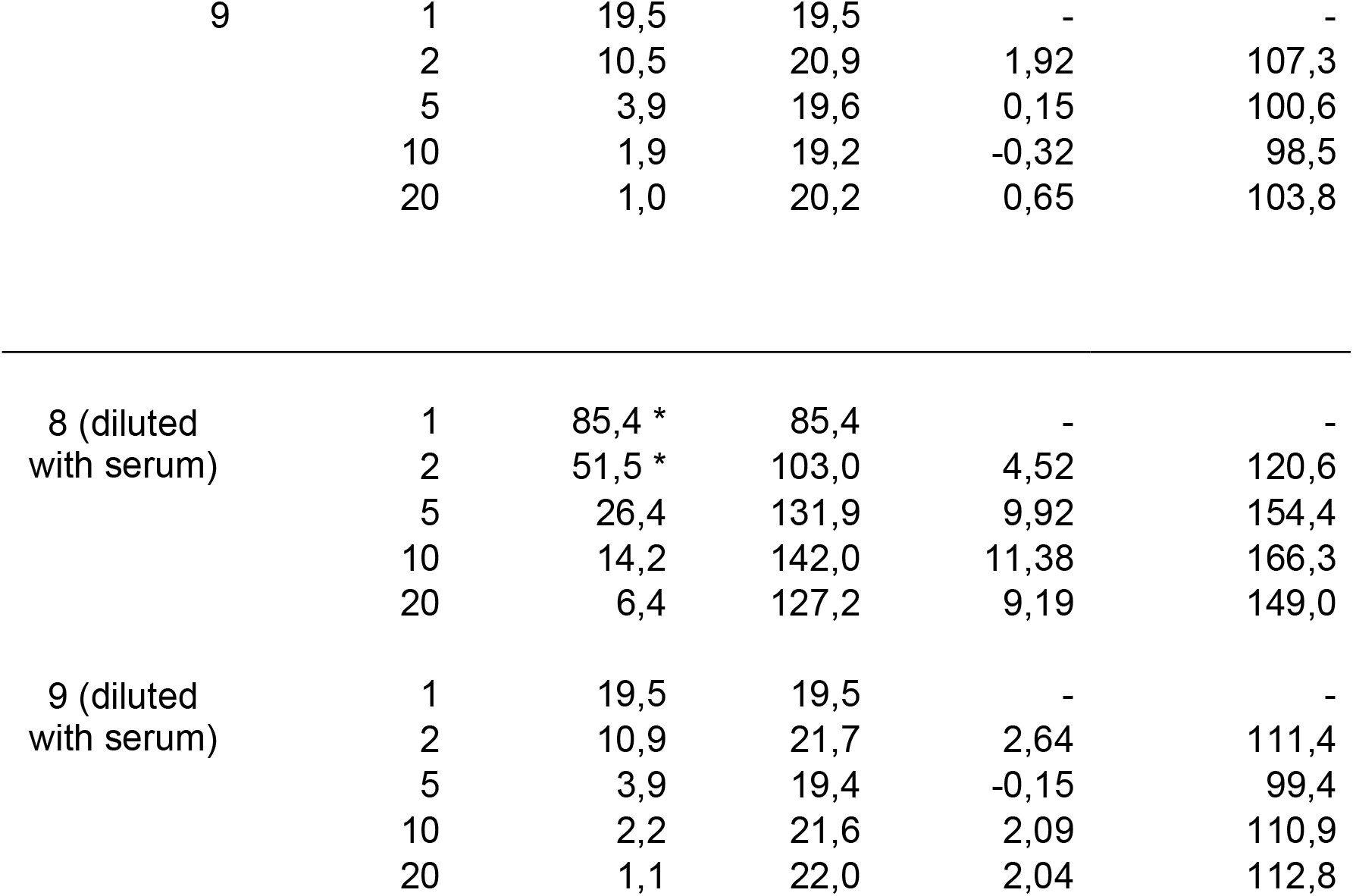
Dilution test with SNIBE method. *The z-score was calculated according Emerson et al (4), and considered acceptable up to 3s. The samples 8 and 9 were diluted also using a negative serum. The asterisk denoted the non-linear sample dilution*.

| Sample | Dilution factor | Measured value | Final value | Z score | Recovery % |
| --- | --- | --- | --- | --- | --- |
| 1 | 1 | 50,1 | 50,1 | - | - |
|  | 2 | 24,2 | 48,4 | -0,94 | 96,6 |
|  | 5 | 9,9 | 49,5 | -0,33 | 98,8 |
|  | 8 | 6,0 | 48,2 | -1,03 | 96,3 |
|  | 10 | 4,5 | 45,5 | -2,52 | 90,4 |
|  | 20 | 2,2 | 44,0 | -2,58 | 87,8 |
| 2 | 1 | 12,0 | 12,0 | - | - |
|  | 2 | 6,6 | 13,2 | 2,69 | 110,0 |
|  | 4 | 2,7 | 10,9 | -2,45 | 90,7 |
|  | 10 | 1,2 | 12,3 | 0,44 | 102,2 |
| 3 | 1 | 59,7 * | 59,7 | - | - |
|  | 5 | 32,7 | 163,4 | 23,84 | 273,7 |
|  | 10 | 15,4 | 153,5 | 21,35 | 257,1 |
|  | 20 | 8,7 | 173,3 | 23,19 | 290,3 |
|  | 50 | 3,0 | 151,6 | 18,30 | 253,9 |
| 4 | 1 | 59,1 * | 59,1 | - | - |
|  | 2 | 35,5 | 71,0 | 4,93 | 120,1 |
|  | 5 | 15,7 | 78,5 | 7,52 | 132,8 |
|  | 10 | 7,1 | 70,8 | 4,84 | 119,8 |
| 5 | 1 | 83,0 * | 83,0 | - | - |
|  | 2 | 51,8 * | 103,6 | 5,93 | 124,8 |
|  | 5 | 28,0 | 140,0 | 13,25 | 168,7 |
|  | 8 | 19,0 | 152,0 | 15,04 | 183,1 |
|  | 10 | 15,5 | 155,0 | 15,45 | 186,7 |
|  | 20 | 7,9 | 158,0 | 15,85 | 190,4 |
|  | 50 | 3,1 | 155,5 | 13,58 | 187,3 |
| 6 | 1 | 49,2 | 49,2 | - | - |
|  | 4 | 13,6 | 54,4 | 2,72 | 110,6 |
|  | 10 | 4,6 | 46,3 | -1,64 | 94,1 |
|  | 20 | 2,5 | 50,2 | 0,52 | 102,1 |
|  | 50 | 0,9 | 45,1 | -1,71 | 91,6 |
| 7 | 1 | 88,1 * | 88,1 | - | - |
|  | 4 | 28,2 | 112,6 | 6,54 | 127,8 |
|  | 10 | 11,5 | 115,3 | 7,15 | 130,9 |
|  | 20 | 5,0 | 100,4 | 3,53 | 114,0 |
| 8 | 1 | 85,4 * | 85,4 | - | - |
|  | 2 | 54,9 * | 109,9 | 6,71 | 128,7 |
|  | 5 | 25,8 | 128,8 | 10,66 | 150,8 |
|  | 10 | 13,0 | 129,5 | 10,79 | 151,6 |
|  | 20 | 6,0 | 120,0 | 8,93 | 140,5 |
| 9 | 1 | 19,5 | 19,5 | - | - |
|  | 2 | 10,5 | 20,9 | 1,92 | 107,3 |
|  | 5 | 3,9 | 19,6 | 0,15 | 100,6 |
|  | 10 | 1,9 | 19,2 | -0,32 | 98,5 |
|  | 20 | 1,0 | 20,2 | 0,65 | 103,8 |
| 8 (diluted<br>with serum) | 1 | 85,4 * | 85,4 | - | - |
|  | 2 | 51,5 * | 103,0 | 4,52 | 120,6 |
|  | 5 | 26,4 | 131,9 | 9,92 | 154,4 |
|  | 10 | 14,2 | 142,0 | 11,38 | 166,3 |
|  | 20 | 6,4 | 127,2 | 9,19 | 149,0 |
| 9 (diluted<br>with serum) | 1 | 19,5 | 19,5 | - | - |
|  | 2 | 10,9 | 21,7 | 2,64 | 111,4 |
|  | 5 | 3,9 | 19,4 | -0,15 | 99,4 |
|  | 10 | 2,2 | 21,6 | 2,09 | 110,9 |
|  | 20 | 1,1 | 22,0 | 2,04 | 112,8 |

## Notes

### Competing Interest Statement

The authors have declared no competing interest.

## References

1 Chen X, Pan Z, Yue S, Yu F, Zhang J, Yang Y et al. Disease severity dictates SARS-CoV-2-specific neutralizing antibody responses in COVID-19. Signal Transduct Target Ther 2020; 5:180. doi: 10.1038/s41392-020-00301-9

2 Lynch KL, Whitman JD, Lacanienta NP, Beckerdite EW, Kastner SA, Shy BR et al. Magnitude and Kinetics of Anti–Severe Acute Respiratory Syndrome Coronavirus 2 Antibody Responses and Their Relationship to Disease Severity. Clin Infect Dis 2021; 72:301–8

3 Wu JT Quantitative immunoassays: a practical guide for assay establishment, troubleshooting and clinical application. AACC Press 2000.

4 Emerson JF, Ngo G, Emerson SS. Screening for interference in immunoassays. Clin Chem 2003; 49:1163–9.

